# Feasibility of remote self-administered cognitive assessment in an Alzheimer’s disease treatment clinic

**DOI:** 10.64898/2026.09.15.26362283

**Authors:** Alyssa N. Kaser, John L. Stricker, Morgan A. Hughes, Nick Corriveau-Lecavalier, Elizabeth A. Boots, Mary M. Machulda, Julie A. Fields, David T. Jones, Stuart J. McCarter, Hugo Botha, Ryan P. Coburn, Camilo Bermudez, Bryan J. Neth, Jonathan Graff-Radford, Vijay K. Ramanan, Nikki H. Stricker

## Abstract

**Introduction:** Remote cognitive assessments offer an accessible, scalable way to support clinical characterization and longitudinal monitoring of cognition. This study evaluates the feasibility of implementing Mayo Test Drive (MTD), a remote, self-administered digital cognitive assessment, within a specialized clinic focused on eligibility assessments for anti-amyloid therapies for Alzheimer’s disease (AD).

**Method:** Two-hundred and nine (*N*=209) patients (M_age_=71.2, 53% female) were evaluated through the Mayo Clinic Rochester Alzheimer’s Disease Treatment Clinic (ADTC) and had the opportunity to complete MTD, either as a remote, patient-initiated session through a patient-facing healthcare portal or during an in-clinic neuropsychological evaluation.

**Results:** Clinical diagnoses included 55.2% mild cognitive impairment (MCI), 31.0% mild dementia, and 11.6% moderate dementia, in addition to 2.2% cognitively unimpaired (CU), as determined by neurology consensus at a multidisciplinary case conference. Among those who initiated MTD (*n*=197/209), 97.5% completed a session (70.6% remotely, 26.9% in clinic). Completion rates were high across diagnostic groups and did not significantly differ by clinical severity (*p*=.12): CU (100%), MCI (99.1%), mild dementia (97.0%), and moderate dementia (89.5%). Remote completion rates overall were high (97.9%). Among completers, administration setting (remote vs. in-clinic) did not significantly differ by clinical stage, χ²(3)=2.37, *p*=.49. Reasons for non-initiation included not receiving or opening portal messages and other/unknown factors.

**Discussion:** Remote digital cognitive assessment is feasible within the context of clinic eligibility evaluations for AD therapies. The high completion rates of MTD across clinical impairment levels in this setting support development of paths for broader use in cognitive characterization in AD care.

## Introduction

The expansion of disease-modifying therapies for Alzheimer’s disease (AD) has increased the need for timely diagnosis and reliable monitoring of cognitive change. At present, anti-amyloid therapies are approved for use in the early symptomatic stages of AD (Cummings et al., 2023; Rabinovici et al., 2025; Ramanan et al., 2023). Optimizing efficacy and safety of these therapies in real-world practice therefore necessitates approaches to efficiently and accurately characterize cognitive status in determining treatment eligibility, and to establish mechanisms for ongoing monitoring once treatment is initiated (Neth et al., 2026). These demands can be time and resource-intensive when applied to real-world populations (Rafii & Aisen, 2025), underscoring the need for efficient, scalable approaches to cognitive assessment.

Comprehensive neuropsychological evaluations are recommended as the best way to objectively assess multi-domain cognition and often are central to clinical staging and characterization in practice (Shaughnessy & Weintraub, 2025). In the context of eligibility evaluations for recently approved AD therapies, neuropsychological assessment can be particularly important for complex or atypical clinical presentations as well as for “borderzone” cases where history and screening tools are insufficient to clarify stage (e.g., cognitively unimpaired versus early mild cognitive impairment). However, access to neuropsychology may not always be available due to long waitlists or other factors, and there are limitations to re-assessing at frequent intervals. In this context, remote and digital cognitive assessments offer a promising complementary approach, with potential advantages in accessibility, scalability, and the ability to support longitudinal cognitive monitoring (Staffaroni et al., 2020).

Prior literature supports the validity and usability of digital cognitive tools in individuals along the AD continuum (Nicosia et al., 2023; Öhman et al., 2021; Patel et al., 2025; Polk et al., 2025). Vanderlip and Stark (2024) suggested that brief digital cognitive assessments may even outperform traditional biomarkers, including cortical amyloid-beta (Aβ) and entorhinal tau, in predicting progression from cognitively unimpaired (CU) status to mild cognitive impairment (MCI). Moreover, digital platforms allow for high-frequency longitudinal monitoring, which can enhance the precision of detecting subtle cognitive change and help track cognitive trajectories over time (Rentz et al., 2016; Samaroo et al., 2020; Thompson et al., 2023). Finally, combining digital cognitive assessments with biomarker and imaging studies may enhance prognostic utility for future cognitive decline (Berron et al., 2024; Tideman et al., 2025). However, less is known about the real-world implementation of remote cognitive assessments within routine AD clinical care. Notably, in the setting of anti-amyloid therapy, scalable cognitive assessments may enhance baseline eligibility determination, support interim monitoring between treatment visits, and enable identification of clinically meaningful change.

Contributing to this effort is Mayo Test Development through Rapid Iteration, Validation, and Expansion (Mayo Test Drive; MTD), a self-administered, multi-device, digital cognitive assessment platform. The MTD cognitive screening battery was designed to assess targeted cognitive domains commonly affected early in AD and other neurodegenerative disorders (Stricker et al., 2022; Stricker at al., 2025) and includes the Stricker Learning Span (SLS), a computer-adaptive verbal word list memory test (Stricker et al., 2022; Stricker et al., 2023), and the Symbols Test, a processing speed and executive function measure that includes visual discrimination demands (Boots et al., 2024; Nicosia et al., 2023). The MTD composite score demonstrates strong alignment with established cognitive composites derived from in-person neuropsychological measures, including the Mayo Preclinical Alzheimer Cognitive Composite (Mayo-PACC; Boots et al., 2024), shows meaningful relationships with AD-related imaging biomarkers (Boots et al., 2024), and good test-retest reliability (Hughes et al., 2026). Prior work has further demonstrated that the SLS performs comparably to the Auditory Verbal Learning Test (AVLT) in distinguishing AD biomarker–defined groups, including amyloid- and tau-positive versus negative individuals, in a largely cognitively unimpaired sample (Stricker, Stricker et al., 2024). These findings highlight the SLS’s sensitivity to early, pathology-related cognitive changes, an important capability for AD treatment clinics (ADTCs) focused on early, accurate identification of treatment-eligible individuals. Recent work examining MTD usability in a large research cohort of adults aged 35 to 100 demonstrated high completion rates for remotely administered sessions (98.5% overall) across both cognitively unimpaired and impaired groups, as well as strong adherence at a 7.5-month follow-up (89%; Patel et al., 2025). A full MTD session is typically completed in approximately 15 to 20 minutes and can be done on multiple device types (e.g., smartphone, tablet, or computer).

Building on this prior work, the present study evaluated the feasibility of implementing MTD clinically within a specialized clinic focused on eligibility assessments for recently approved AD therapies. Specifically, we examined implementation outcomes, including rates of test initiation and completion, factors contributing to non-initiation, and whether completion rates, method of administration (remote self-initiated versus in clinic), and session duration differed across levels of clinical severity. This work aimed to clarify whether self-administered digital cognitive assessment is feasible for patients referred for anti-amyloid therapy evaluations in real-world AD treatment settings.

## Methods

### Participants

Patients included in this study were evaluated through the Mayo Clinic Rochester Alzheimer’s Disease Treatment Clinic, a multidisciplinary clinical program designed to assess eligibility for anti-amyloid therapies for AD. The ADTC evaluation integrates neurology, neuropsychology, neuroimaging, laboratory studies, and care partner input to support diagnostic clarification and treatment planning. Clinical diagnoses and syndromic classifications were determined via neurologic diagnosis derived from multidisciplinary case conference review, and included CU, MCI, mild dementia, and moderate dementia. The ADTC clinical workflow and evaluation framework have been described in detail elsewhere (Neth et al., 2026).

### Human Research and Informed Consent

This retrospective study received exempt determination from the Mayo Clinic Institutional Review Board under 45 CFR 46.104(d), category 4(iii). Consistent with the Minnesota Research Authorization Policy, individuals who declined use of their medical records for research purposes were excluded from analyses.

### Mayo Test Drive Procedures and Workflow

Within the ADTC workflow, patients are first provided the opportunity to complete MTD remotely via a personalized patient portal message sent approximately eight days prior to their in-person neuropsychological evaluation. The message provides brief instructions and a unique link that directly opens a web-based test session. A quick response (QR) code version of the link is also provided to allow device choice flexibility. Patients are instructed to complete the assessment independently in a single uninterrupted sitting using a smartphone, tablet, or personal computer, allowing approximately 15-20 minutes in a quiet, distraction-free environment. Instructions specify that assistance is permitted only for accessing the testing website or resolving technical issues. The web browser should not be closed during testing, and testing links should not be shared or used by anyone else. Patients who initiate but do not complete MTD remotely, as well as those who do not engage with the portal message, are subsequently offered in-clinic administration of MTD during their neuropsychological evaluation, when applicable. For in-clinic administration, patients typically complete MTD on a tablet device at the conclusion of neuropsychological testing. Psychometrists provide the device and facilitate access but typically do not provide further assistance during the session. Figure 1 illustrates the ADTC clinical workflow and corresponding MTD initiation and completion pathways across remote and in-clinic administration methods.

**Figure 1.**
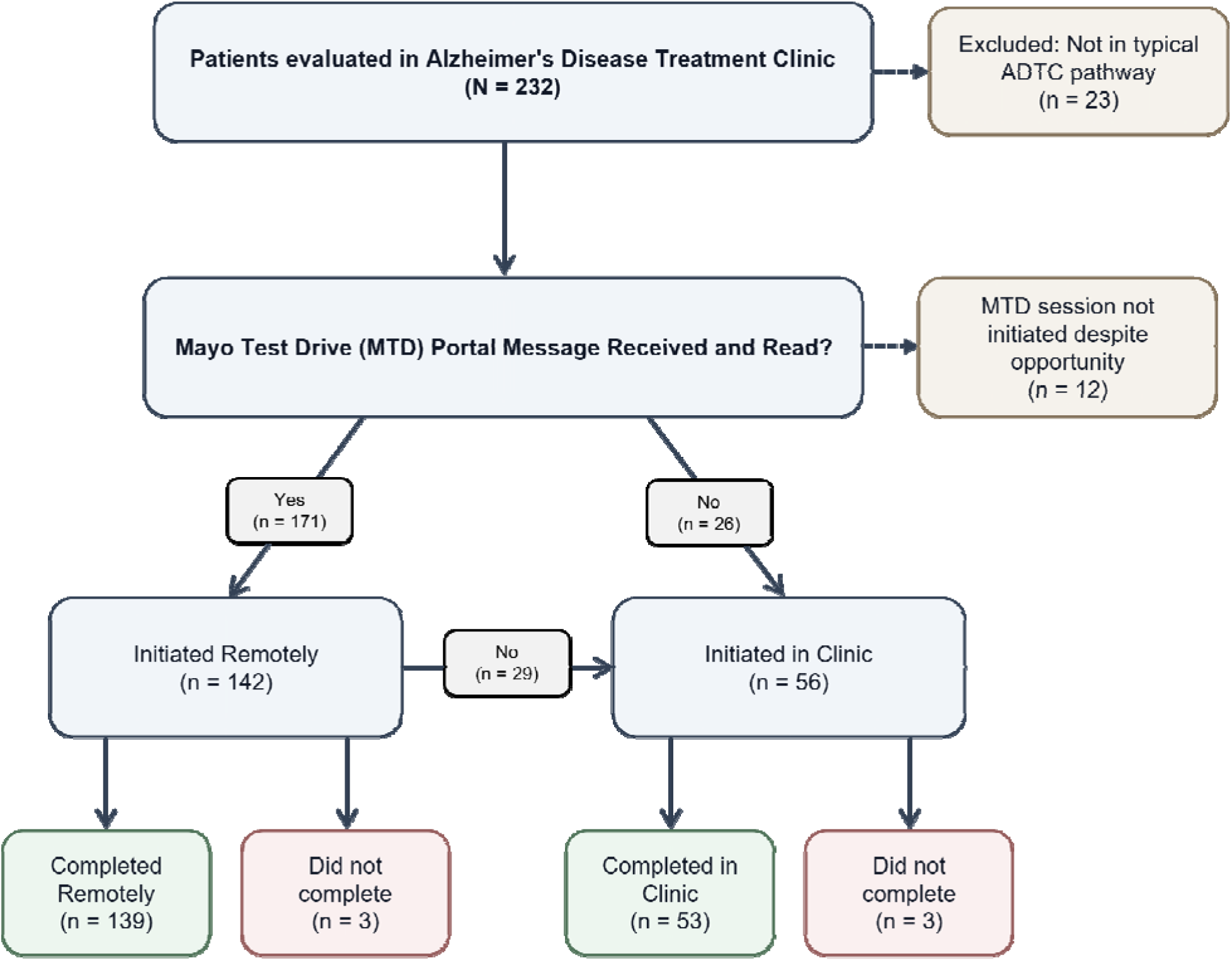
Clinical workflow and completion pathways for Mayo Test Drive administration in an Alzheimer’s Disease Treatment Clinic. *Note*. One patient who initiated but did not complete MTD remotely subsequently completed a session in clinic.

### MTD Initiation and Completion

Initiation and completion thresholds were defined following terminology and procedures outlined by Patel et al. (2025). In this framework, initiation of an MTD session refers to the point at which a patient opens the testing platform and begins the session, operationalized as making a response on the location-selection screen that appears after the welcome instructions. Completion was defined as finishing an MTD session after it had been initiated. For patients who did not initiate a remote MTD session, reasons for non-initiation were categorized based on medical record review and platform metadata. These factors included inactive patient portals, failure to receive or open patient portal messages, and other or unknown factors. Among patients who initiated an MTD session, completion rates were examined overall and further stratified by clinical diagnosis and method of administration (remote versus in-clinic).

### Statistical Analysis

Descriptive statistics were generated to characterize demographic and clinical features of the cohort, as well as rates of MTD initiation and completion. Chi-square or exact tests were conducted to examine differences in MTD completion rates and administration methods by clinical severity, as appropriate based on expected cell counts. Differences in session duration across clinical severity levels were examined using a Kruskal–Wallis test, with Dunn’s pairwise comparisons and Bonferroni correction for multiple comparisons. Differences in session duration between remote and in-clinic administration were examined using the Mann–Whitney U test. Statistical analyses were performed using SPSS (version 29.0, SPSS, Inc., Chicago, IL).

## Results

### Participants

Between October 2, 2023 and December 31, 2024, 232 patients were evaluated in the ADTC; we chose this time range to maintain consistency with the patients presented in our recently published manuscript that provides in-depth characterization of this cohort (Neth et al., 2026). Of these, 23 patients did not follow the typical ADTC clinical pathway and therefore were not offered MTD, resulting in exclusion from the feasibility analyses conducted in this work.

Among the 209 patients who were provided with the opportunity to complete MTD (either remotely via receipt of a personalized patient portal link for remote completion or in-clinic during their neuropsychological evaluation), 12 of these patients received an MTD portal message but did not initiate a session remotely or in-clinic. Specifically, nine were not seen for an ADTC-specified neuropsychological evaluation, precluding in-clinic administration. The neuropsychologist decided not to administer MTD in one patient due to severe visuospatial processing impairments related to a posterior cortical atrophy phenotype. An MTD session was planned in clinic for two patients but did not meet criteria for objective task initiation, defined as making a response on the location-selection screen. In both cases, the psychometrist determined that the patients were unable to read and follow the initial prompt to select a location prior to the administration of any formal test instructions. Among the nine patients not seen in an ADTC neuropsychological appointment slot, six patients opened the MTD patient portal message but did not initiate a remote session and subsequently did not receive MTD in clinic, as they did not have an in-person neuropsychological visit during which MTD could be administered. A total of 197 patients initiated an MTD session either remotely or in clinic and were included in the primary analyses examining MTD initiation, completion, and administration rates. All 209 participants were additionally included in select secondary analyses for descriptive purposes.

### Sample Characteristics

Demographic characteristics of the 197 patients included in the primary analyses are described in Table 1. Baseline cognitive screening using the Kokmen Short Test of Mental Status (STMS) indicated a mean score of 28.63 (SD = 5.93, range = 4 – 38) out of 38 possible (Kokmen et al., 1991). Informant-reported functional abilities, as measured by the Functional Activities Questionnaire (FAQ; Pfeffer et al., 1982), reflected a mean score of 7.14 (SD = 6.13, range = 0 – 29), with higher scores reflecting greater impairment in instrumental activities of daily living. The sample had a mean Clinical Dementia Rating (Morris, 1993) Sum of Boxes (CDR-SB) score of 2.58 (SD = 1.96, range = 0 – 10), with the following distribution of CDR global scores: 0 (3.8%), 0.5 (63.9%), 1 (29.3%), and 2 (3.0%). Neurology clinical diagnosis determined via multidisciplinary case conference review classified 2.5% of patients as CU, 54.3% as having MCI, 33.5% as having mild dementia, and 9.6% as having moderate dementia. APOE 4 status was available for most participants: 118 (59.9%) were 4 positive, 61 (30.9%) were 4 negative, and 18 (9.1%) had not been tested. Amyloid status, determined by amyloid positron emission tomography (PET) or cerebrospinal fluid (CSF) biomarkers through lumbar puncture, was positive in 164 participants (83.2%), negative in 22 participants (11.2%), and not tested in 10 participants (5.1%). Among the 10 participants who did not undergo amyloid testing, several were not reviewed for treatment eligibility because they were cognitively unimpaired (n = 1), had moderate dementia (n = 4), or had other factors that would preclude treatment consideration, including suspected non-AD etiologies or exclusionary imaging findings.

**Table 1.** Sample Characteristics for Patients who Initiated an MTD Session.

| **Characteristic** | **Sample (N = 197)** |
| --- | --- |
| Age, *M (SD)* | 71.1 (7.1) |
| Sex, n (%) Female | 105 (53.3) |
| Race, n (%) |  |
| White/European American | 189 (95.9) |
| Black/African American | 3 (1.5) |
| Asian | 2 (1.0) |
| Native American | 1 (0.5) |
| Undisclosed | 2 (1.0) |
| Ethnicity, n (%) Hispanic | 6 (3.0) |
| Education, *M (SD)* | 15.9 (2.4) |
| ADTC Referral Source, n (%) Internal | 115 (58.4) |
| APOE ε4 Status, n (%) | |
| ε4 positive | 118 (59.9) |
| ε4 negative | 61 (30.9) |
| Not available | 18 (9.1) |
| Amyloid Status, n (%)<sup>1</sup> |  |
| Positive | 164 (83.2) |
| Negative | 22 (11.2) |
| Not available | 10 (5.1) |
| STMS, *M (SD)* | 28.6 (5.9) |
| CDR-SB, *M (SD)*<sup>2</sup> | 2.6 (2.0) |
| CDR-Global, n (%)<sup>2</sup> |  |
| 0 | 5 (3.8) |
| 0.5 | 85 (63.9) |
| 1 | 39 (29.3) |
| 2 | 4 (3.0) |
| FAQ, *M (SD)*<sup>2</sup> | 7.1 (6.1) |
| Clinical diagnosis, n (%) |  |
| Cognitively Unimpaired | 5 (2.5) |
| Mild Cognitive Impairment | 107 (54.3) |
| Mild Dementia | 66 (33.5) |
| Moderate Dementia | 19 (9.6) |

*Note.* MTD = Mayo Test Drive; APOE = apolipoprotein E; STMS = Kokmen Short Test of Mental Status (total possible score of 38); CDR-SB = Clinical Dementia Rating – Sum of Boxes; CDR-Global = Clinical Dementia Rating – Global; FAQ = Functional Activities Questionnaire; Amyloid status determined by amyloid PET or CSF biomarker studies; <sup>1</sup>Regionally positive, globally negative n=1; <sup>2</sup>Based on available data; Missing observations were noted for CDR-SB (n=67), CDR-Global (n=64), and FAQ (n=36).

### MTD Initiation and Completion Rates

Of those who initiated MTD, 72.1% (142/197) initiated remotely and 28.4% (56/197) initiated in clinic. A total of 171 individuals opened and read their portal message, of whom 142 initiated a remote session (83.0%; 142/171). Overall, 56 individuals initiated an MTD session in clinic, including those who did not open or read the portal message (n = 26), those who read the message but did not initiate a remote session (n = 29), and one individual who initiated in clinic following an incomplete remote session (n = 1; Figure 1). Among the 197 patients who initiated an MTD session either remotely or in clinic, 97.5% successfully completed a session.

Remote completion rates were high once MTD was initiated, with 97.9% (139/142) of patients completing a remote session. Among participants who completed MTD remotely, 52.5% used a desktop or laptop computer, 25.9% used a smartphone, 19.4% used a tablet, and 2.2% selected “other/not sure.”

Similarly, in-clinic completion rates were also high, with 94.6% (53/56) completing a session in clinic once a session was initiated. MTD was completed via tablet for most in-clinic sessions (75.5%). An additional 15.1% completed MTD via smartphone, 1.9% selected computer/laptop, and 7.5% selected “other/not sure.” Table 2 outlines MTD initiation and completion rates by administration pathway.

**Table 2.** MTD Initiation and Completion Rates by Administration Pathway.

| **MTD Subgroup** | **Completed Sessions (n/N)** | **Completion Rate (%)** | **Interpretation** |
| --- | --- | --- | --- |
| All patients who were offered MTD | 192 / 209 | 91.9 | Most who had an opportunity for MTD initiated and completed a session |
| All patients who initiated MTD | 192 / 197 | 97.5 | Completion was high among patients who initiated an MTD session |
| Patients who opened and read MTD portal message | 139 / 171 | 81.3 | Most who engaged with the portal message completed MTD remotely, though some did not initiate a session |
| Initiated remotely | 139 / 142 | 97.9 | Nearly all patients who initiated MTD remotely completed a session |
| Initiated in clinic | 53 / 56 | 94.6 | Completion remained high when MTD was initiated in clinic |

### MTD Completion and Administration by Clinical Severity

Collapsing across remote and in-clinic settings, MTD completion rates did not significantly differ by clinical severity (Fisher–Freeman–Halton exact test, *p* = .119). Completion rates were high across diagnostic groups (Table 3), including CU (100%), MCI (99.1%), mild dementia (97.0%), and moderate dementia (89.5%). The proportion of remote versus in-clinic administration also did not significantly differ by clinical severity, χ²(3) = 2.37, *p* = .49. Sessions were done remotely in 80.0% (4/5) of CU individuals, 76.4% (81/106) of those with MCI, 67.2% (43/64) of those with mild dementia, and 64.7% (11/17) of those with moderate dementia.

**Table 3.**
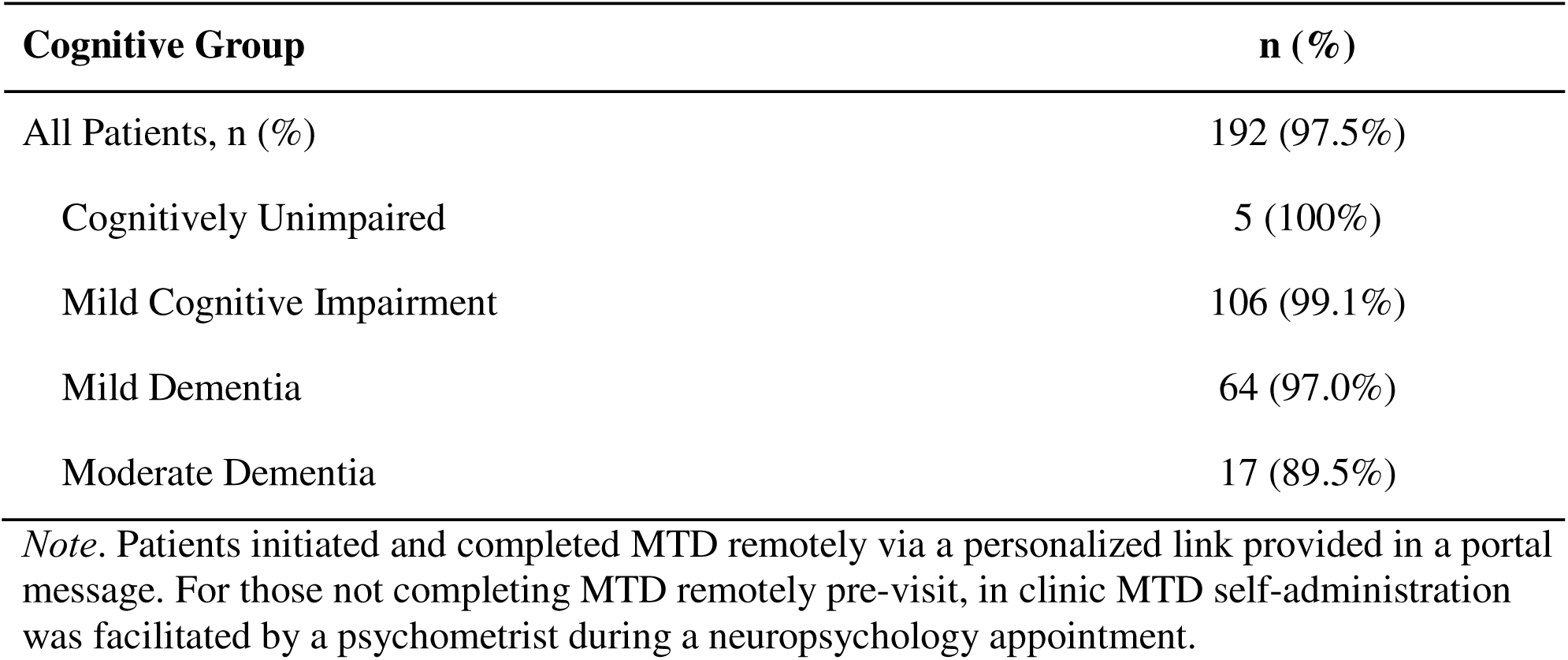
MTD Completion Rates for Patients who Initiated an MTD Session (n=197) by Cognitive Group.

### MTD Session Duration by Clinical Severity

MTD session duration significantly differed across clinical severity levels, *H*(3) = 21.17, *p* < .001. Median session duration was 20.0 minutes (IQR = 6.0) among CU participants, 18.0 minutes (IQR = 6.0) among those with MCI, 19.5 minutes (IQR = 9.0) among those with mild dementia, and 30.0 minutes (IQR = 29.0) among those with moderate dementia. Dunn’s post-hoc comparisons with Bonferroni correction indicated that individuals with moderate dementia had significantly longer session durations than individuals with MCI (adjusted *p* < .001) and mild dementia (adjusted *p* = .02). No other pairwise comparisons were statistically significant. MTD session duration also significantly differed by administration method, with remote sessions demonstrating longer median durations than in-clinic sessions (20.0 vs. 17.0 minutes; IQR = 8.0 for both), U = 4,880.50, *z* = 3.58, *p* < .001.

### Reasons for Non-Initiation of Remote MTD Session

Among patients who did not initiate a remote MTD session (n = 55; 27.9% of analytic sample), barriers included inactive patient portals (2.0%), portal messages not received (4.1%), portal messages delivered but not read (9.6%), and other or unknown factors (12.2%; Table 4). Documented “other” factors included logistical constraints, such as the patient being unavailable between receipt of the portal message and the in-clinic evaluation.

**Table 4.** Reasons for Non-Initiation of Remote Mayo Test Drive Session.

| **Factors** | **n (%)** |
| --- | --- |
| Inactive patient portal | 4 (2.0%) |
| Portal message not sent | 8 (4.1%) |
| Portal message delivered but not read | 19 (9.6%) |
| Other<sup>1</sup> / unknown factors | 24 (12.2%) |
| **Total** | **55 (27.9%)** |

*Note.* Patients reflected in this table are those who did not initiate MTD remotely and subsequently initiated MTD in clinic (n=55); <sup>1</sup> Other documented factor reflects logistical constraints (e.g., patient being unavailable from time of portal message received to time of in-clinic evaluation).

## Discussion

The present study evaluated the feasibility of clinically implementing a self-administered digital cognitive assessment screening battery within the Mayo Clinic Rochester ADTC, a specialized clinic providing eligibility assessments for anti-amyloid therapy for AD. The study focused on key implementation outcomes, including rates of MTD initiation and completion, barriers to engagement, and whether administration methods or completion rates differed across severity of cognitive impairment. Overall, findings demonstrated high rates of MTD initiation and completion across a clinical sample of individuals ranging from CU to those with moderate dementia, supporting the practical feasibility of integrating remote digital cognitive assessment into AD clinical workflows.

Among patients who initiated MTD, nearly all successfully completed a session (97.5%), with similarly high completion rates for both remote (97.9%) and in-clinic (94.6%) administration. Importantly, most patients who opened and viewed the patient portal message subsequently initiated remote testing (83%), suggesting strong patient willingness to engage with self-administered digital assessment when it is part of the clinical workflow. These findings extend prior research demonstrating high usability and completion rates of self-administered remote cognitive assessments in research settings (Dhanam et al., 2026; Edgar et al., 2021; Patel et al., 2025; Thompson et al., 2024) and suggest that comparable levels of engagement can be achieved in clinical care settings.

Completion rates remained high among patients with mild (97%) and moderate (89.5%) dementia, suggesting that individuals with greater cognitive impairment can successfully engage with self-administered digital assessment platforms within this clinical context. Notably, most patients with mild and moderate dementia completed MTD remotely (67.2% and 64.7%, respectively), further supporting the accessibility of digital cognitive assessment across a spectrum of cognitive functioning (Ohman et al., 2021). At the same time, a small subset of patients (*n* = 3) were unable to initiate or complete MTD due to cognitive barriers, including difficulty understanding instructions or severe visuospatial impairment, highlighting important limitations of fully self-administered approaches. These findings suggest that while remote digital tools may be feasible for many individuals with cognitive impairment, some patients with more advanced disease or sensory/motor deficits may require additional support or alternative assessment approaches, although this limitation may be less relevant for amyloid-reducing therapies, which are intended for patients with mildly symptomatic AD. This observation aligns with recommendations from the Global CEO Initiative on AD (CEOi) on clinical adoption of digital cognitive assessment, which note that cognitive symptoms themselves may introduce barriers to successful remote assessment (Thompson et al., 2025).

Although completion rates did not differ significantly by clinical severity, session duration varied across groups. Patients with moderate dementia had longer session durations than those with MCI and mild dementia, whereas durations were relatively similar among CU, MCI, and mild dementia groups. Session duration also differed significantly by administration method, with remote sessions demonstrating longer median durations than in-clinic sessions. Several factors may contribute to this variability. First, MTD does not impose time-out limits, allowing patients to take as much time as needed to respond to individual tasks. As a result, session duration may more fully capture differences in response time and pacing than assessments with fixed time limits. Administration method may also influence session duration. Remote administration provides greater flexibility in when the assessment is completed, whereas in-clinic administration occurs within a more structured testing environment, with psychometrists facilitating access to the assessment during a scheduled clinical visit. This built-in structure may reduce opportunities to pause or otherwise interrupt testing, potentially contributing to shorter in-clinic session durations. Thus, session duration likely reflects a combination of cognitive factors and contextual factors related to the testing environment, administration workflow and participant behavior during the test session.

This work contributes to the growing literature on digital cognitive assessment by extending feasibility findings to clinical care, as much of the existing literature evaluating digital cognitive tools in AD has been conducted in research cohorts. Comparatively fewer studies have examined implementation and feasibility within real-world clinical care settings. Recent work by Fowler et al. (2025) demonstrated that in-clinic digital cognitive assessments for ADRD can be feasibly integrated into primary care-based dementia screening workflows, with relatively low refusal rates among patients (22%). Additionally, they identified implementation barriers, such as variability in provider engagement and competing clinical demands, that will inform future implementation efforts. The current findings build upon this work by demonstrating successful implementation within a specialty AD treatment model, where cognitive characterization plays an important role in treatment eligibility determination and ongoing clinical management.

The present study also underscores the importance of workflow-related factors in successful implementation of remote digital cognitive assessment. Many instances of remote non-initiation appeared attributable to logistical barriers rather than difficulties completing the assessment itself, including inactive patient portals, unread portal messages, and an absent MTD clinical order. These findings suggest that patient engagement with digital assessment may be strongly influenced by healthcare system infrastructure and patient communication processes. Future implementation efforts may benefit from more standardized ordering procedures, automated reminders, and additional outreach methods for individuals who do not routinely use electronic patient portals. Notably, the current ADTC clinical workflow allowed many patients who did not complete MTD remotely to subsequently complete testing during their in-clinic neuropsychological evaluation, thereby maximizing overall completion rates and accessibility. This hybrid approach may represent a practical model for integrating remote digital cognitive assessment into specialty memory care settings.

The current findings should be considered in the context of several limitations. First, the sample was drawn from a single tertiary academic medical center with a predominantly White, non-Hispanic, highly educated patient population, potentially limiting generalizability to more diverse populations. Differences in technological access, digital literacy, and healthcare infrastructure may influence feasibility in other clinical settings. Additionally, we did not capture whether patients received assistance with portal communication or remote MTD initiation, or whether a care partner was already involved in managing the patient’s electronic patient portal. Thus, we cannot determine the extent to which informal assistance may have contributed to successful remote engagement. Future work may explore this, in addition to approaches for involving care partners, when appropriate, to facilitate communication and session initiation while maintaining the self-administered nature of the assessment. Second, patients evaluated within a specialty AD treatment clinic may be particularly motivated to engage in diagnostic and treatment-related procedures, which could also contribute to higher participation rates than might be observed in other clinical or research settings, such as primary care. Third, this study focused specifically on implementation feasibility rather than diagnostic accuracy, longitudinal sensitivity to cognitive change, or clinical utility of MTD-derived data. Future studies examining diagnostic utility and longitudinal cognitive monitoring in those initiating anti-amyloid therapies are planned from the Mayo Clinic Rochester ADTC cohort and will be important for clarifying the broader clinical utility of remote digital cognitive assessments in AD care. Finally, because patients who did not initiate testing remotely were subsequently offered in-clinic administration, the current findings may underestimate barriers that could emerge in a fully remote implementation model.

## Conclusion

As AD treatment models continue to evolve with the expansion of disease-modifying therapies, efficient and scalable approaches to cognitive assessment are increasingly needed. The present study demonstrates that MTD, a remote digital cognitive assessment platform, can be successfully implemented within a real-world ADTC, with high rates of initiation and completion across a range of cognitive impairment severity. Findings suggest that remote digital cognitive assessment is feasible for many patients evaluated for AD treatment and may serve as a practical complement to traditional neuropsychological assessment. Future work is needed to examine longitudinal implementation and the clinical utility of MTD for supporting ongoing monitoring in patients receiving anti-amyloid therapies for AD.

## Highlights

- Remote cognitive assessment was feasible in an Alzheimer’s disease treatment clinic
- Mayo Test Drive achieved 97.5% completion after session initiation
- Remote testing was completed across MCI and mild and moderate dementia severity levels
- Clinical workflow barriers accounted for most remote non-initiation

## Data Availability

All data produced in the present study are available upon reasonable request to the authors.

## Funding and Acknowledgments

Research reported in this publication was supported by the National Institute on Aging of the National Institutes of Health under Award Numbers R01AG081955, R21 AG073967, P30 AG062677, U01 AG006786, and R01 AG034676 (the Rochester Epidemiology Project). This work was also supported by the Kevin Merszei Career Development Award in Neurodegenerative Diseases Research IHO Janet Vittone, MD, the GHR Foundation, and the Mayo Foundation for Medical Education and Research. The content is solely the responsibility of the authors and does not necessarily represent the official views of the National Institutes of Health or other sponsors. A Mayo Clinic invention disclosure has been submitted for the Stricker Learning Span and the Mayo Test Drive platform (NHS, JLS). We have no other conflicts of interest to disclose related to this work.

## Declaration of Competing Interests

ANK declares no competing interests.

JLS reports grants from NIH during the conduct of the study, is a co-founder and shareholder of Cephlodyne Neurotechnologies, and is a named inventor on a Mayo Clinic invention disclosure related to the Stricker Learning Span and the Mayo Test Drive platform. JLS receives no personal compensation from any commercial entity.

NCL reports funding from the Brain & Behavior Research Foundation during the conduct of the study and receives no personal compensation from any commercial entity.

EAB declares no competing interests.

MMM reports grants from NIH during the conduct of the study.

JAF reports grant funding from the NIH during the conduct of the study.

DTJ receives support from the NIH, is co-founder and shareholder of Cephlodyne Neurotechnologies, and is named inventor on patents assigned to Mayo Clinic related to neuroimaging analysis.

SJM receives support from the American Academy of Sleep Medicine foundation and NIH. He serves as an investigator for a clinical trial sponsored by Cognition Therapeutics but does not receive personal compensation. He served as a consultant for Open Evidence.

HB reports grants from NIH during the conduct of the study. He also receives research funding from Cervomed as site PI for NCT07033481.

RPC declares no competing interests.

CBN reports no financial disclosures or conflicts of interest.

JGR reports grants from NIH during the conduct of the study. He reports serving on the Data and Safety Monitoring Board for StrokeNET NINDS, serves as site investigator for trials sponsored by Eisai and cognition therapeutics, serves as a consultant to OpenEvidence, and reports honoraria for serving as faculty member for American Academy of Neurology and IMPACT AD clinical trials course, outside the submitted work.

BJN declares no competing interests.

VKR reports receiving research funding from the NIH, the Kogod Center for Aging, and the Mangurian Foundation for Lewy Body disease research; has provided CME and other educational content for Medscape, Expert Perspectives in Alzheimer’s Disease, Clinical Care Options, PeerView Institute (CME activity supported by an educational grant from Eli Lilly), and the Association of Diagnostic and Laboratory Medicine (webinar supported by an educational grant from Roche); has received speaker and conference session honoraria from the American Academy of Neurology Institute; has served on and chaired a Data Safety Monitoring Board for a clinical trial supported by the Weston Family Foundation; is PI for a clinical trial supported by the Alzheimer’s Association; is site Co-PI for the Alzheimer’s Clinical Trials Consortium; and is a site clinician for clinical trials supported by Eisai, Cognition Therapeutics, the Alzheimer’s Treatment and Research Institute at USC, and Transposon Therapeutics, Inc.

NHS reports grants from NIH during the conduct of the study. NHS reports grant support from the Kevin Merszei Career Development Award in Neurodegenerative Diseases Research IHO Janet Vittone, MD (Mayo Clinic Center for Clinical and Translational Science) during the conduct of the study. NHS reports consulting fees from the University of Georgia, outside the submitted work. NHS is a named inventor on a Mayo Clinic invention disclosure related to the Stricker Learning Span and the Mayo Test Drive platform. NHS receives no personal compensation from any commercial entity.

## Notes

### Author Declarations

Institutional Review Board under 45 CFR 46.104(d), category 4(iii) of Mayo Clinic gave ethical approval for this work.

